# Prevalence and sociodemographic factors associated with unwanted pregnancy among adolescent girls in Kakuma Refugee Camp, Kenya: a cross-sectional mixed-methods study

**DOI:** 10.64898/2026.09.14.26363074

**Authors:** Benson Simba, Louisa Ndunyu, Lilian Ogonda

**Affiliations:** Department of Public Health, School of Public Health Maseno University, Private Bag, Maseno Kenya; Department of Biomedical Science and Technology, School of Public Health Maseno University, Private Bag, Maseno Kenya

**Keywords:** adolescent girls, unwanted pregnancy, refugee settings, sociodemographic factors, Kenya, Prevalence, adolescents, pregnancy, determinants and sociodemographic

## Abstract

Globally, an estimated 21 million adolescent girls aged 15–19 years become pregnant annually, with the burden often higher in refugee contexts. Adolescent pregnancy remains an important public health concern, particularly in humanitarian and refugee settings. Despite the availability of sexual and reproductive health services in Kakuma Refugee Camp, unwanted pregnancy among adolescent girls persists. According to routine health data from the United Nations High Commissioner for Refugees (UNHCR), 1,079 adolescent pregnancy cases were reported between January 2024 and July 2025 in Kakuma Refugee Camp. However, evidence on the factors associated with unwanted pregnancy among adolescent girls in this multinational refugee setting remains limited. This study estimated the prevalence of unwanted pregnancy and examined socio-demographic factors associated with unwanted pregnancy among adolescent girls in Kakuma Refugee Camp.

We conducted a community-based analytical cross-sectional mixed-methods study among 431 adolescent girls aged 12–18 years recruited through multistage cluster sampling. Quantitative data were collected using a pretested structured questionnaire. For the qualitative data, audio-recorded interviews were conducted with four purposively selected health workers as key informants, together with a focus group discussion involving adolescent girls. Quantitative data were analysed using descriptive statistics, chi-square tests, bivariable logistic regression to estimate crude odds ratios (CORs), and multivariable logistic regression to estimate adjusted odds ratios (AORs), with statistical significance set at p ≤ 0.05. Qualitative data were analysed thematically.

The prevalence of unwanted pregnancy was 28.7%. The proportion varied considerably across camp sections, with the highest prevalence observed in Kakuma 2 (56.7%) and Kakuma 4 (54.1%). In adjusted analysis, adolescents aged 12–14 years had lower odds of unwanted pregnancy than those aged 15–18 years, although the association was borderline statistically significant (AOR = 0.348; 95% CI: 0.121–1.000; p = 0.050). Married adolescents had substantially higher adjusted odds of unwanted pregnancy than single/divorced/separated adolescents (AOR = 5.108; 95% CI: 3.065–8.512; p = 0.001), and Christian adolescents had higher adjusted odds than adolescents of other religions (AOR = 3.010; 95% CI: 1.087–8.337; p = 0.034). Compared with Kakuma 4, lower adjusted odds were observed in Kakuma 1 (AOR = 0.168; 95% CI: 0.059–0.483; p < 0.001) and Kakuma 3 (AOR = 0.073; 95% CI: 0.021–0.248; p < 0.001), while Kakuma 2 did not differ significantly. Qualitative findings highlighted older adolescent age, being out of school, lower educational attainment, and socioeconomic vulnerability as factors perceived to increase vulnerability to unwanted pregnancy.

Unwanted pregnancy remains an important public health concern among adolescent girls in Kakuma Refugee Camp. The findings identify marital status, religion, and camp location as important independent correlates, with age showing borderline evidence of an adjusted association. School attendance and education level were associated with unwanted pregnancy in unadjusted analyses but were not independently significant in the adjusted results presented. These findings support context-specific interventions that improve contraceptive access, support school retention, engage married adolescents, and address social and structural vulnerabilities across Kakuma camp sections.

## Introduction

Globally, an estimated 21 million adolescent girls aged 15–19 years become pregnant annually, nearly half of which are unintended, and a substantial proportion result in unsafe abortion, particularly in low-resource settings [1]. In humanitarian and refugee contexts, the burden is often higher due to compounded vulnerabilities, including disrupted health systems and limited access to sexual and reproductive health (SRH) services [2]. Evidence suggests that adolescent pregnancy prevalence ranges from 21% to over 40% in refugee camps, reflecting compounded vulnerabilities associated with displacement [3].

Unwanted pregnancy among refugee adolescent girls represents a major public health burden [1] These pregnancies contribute substantially to morbidity and mortality among adolescent girls, accounting for up to 23% of the disease burden in this age group, and are associated with poor maternal and neonatal outcomes, unsafe abortions, school dropout, and long-term socio-economic disadvantage [4]. Routine health data indicates 1,079 adolescent pregnancies recorded between January 2024 and July 2025 Kakuma camp [2]. Although adolescent specific maternal mortality estimates for Kakuma Refugee Camp are not readily available, pregnancy and childbirth-related complications remain among the leading causes of mortality among girls aged 15–19 years globally [1] Adolescents may face heightened risks in humanitarian settings where displacement and barriers to timely, comprehensive maternal and reproductive health services can compound pregnancy-related vulnerabilities

An unwanted pregnancy is a pregnancy that is reported to have occurred when no children, or no more children, were desired [5]. Whilst early and forced marriages certainly occur out-side conflict affected regions, such marriages within conflict affected regions have additional dimensions and complexities [6]. In Refugee settings birth rates have been estimated to be four times higher among refugee adolescents than in the rest of the populations [7].

Demographic factors have been found to influence unwanted pregnancy. In a Tanzania refugee study gender and age of young women were found to increase the risk of sexual violence and other poor SRH outcomes [6]. Populations in which age at first intercourse is low tend to have early childbearing and subsequently give birth to more children, leading to high fertility rates which increased unwanted pregnancy [8]. Attempts to address early marriages in Kakuma have yielded minimal results with one in three girls getting married before the age of 18 [9].

Age of household head and sex of household head have been statistically associated with adolescent pregnancy. Movement of refugee women from one country to another may lead to separation of sexual partners [10]. Conflict can increase the protective role of families as young women tend to be at higher risk of rape, sexual exploitation and abuse when cut off from family structures. Absence of males from the household, through conflict mortality, imprisonment and military membership, can leave households vulnerable to poverty and result in the engagement of females in economic activities which increase the risk of poor SRH [11]. This study is designed to explore how this practice has changed over time and how it is influencing unwanted pregnancy outcomes in Kakuma.

Factors such as occupation influence unwanted pregnancy [12]. Conflict and natural disasters push individuals, and sometimes entire communities, into poverty as crises destroy livelihoods, result in the loss of property and separate people from their economic networks [13]. In Kakuma Camp, Women in the camp are less likely to be entrepreneurs than men, and their businesses are more likely to be informal and have less invested in them [14].

In refugee settings, access to resources is often mediated through household structures, with the characteristics and control of the household head significantly influencing vulnerability, leaving individuals particularly women and those without strong family support at heightened risk of marginalization and limited access to livelihood assets [15]. In Kakuma for instance food assistance to most vulnerable households is at 57% leaving other vulnerable households to seek alternative coping mechanisms [14]. Despite ongoing SRH interventions, Edmond et al 2001 found out that some male adolescents are involved in sex with female adults in exchange for food or money hence the need for more insights as well as understand changes over time [16].

Accessing education, through schools, has been identified as a key determinant and protective factor in relation to most measures of SRH [17]. Education for girls’ delays marriage and reduces desired family size due to aspirations of better standard of living and increased opportunity cost of each child born [18]. Studies show that more educated males in refugee settings are more positive about the use of contraceptives compared to uneducated men [19]. Recent evidence indicates that educational deprivation remains high among female refugees in Kakuma, with approximately 66% of women having no formal education, highlighting persistent structural barriers to girls’ education in the camp [2]. Educational access is an important protective factor for adolescent sexual and reproductive health, yet educational deprivation remains substantial among female refugees in Kakuma. The relationship between school participation and unwanted pregnancy among adolescent girls in the camp, however, remains insufficiently characterized.

Religion has been shown to have an association with unwanted pregnancy. A study done in Kakuma to explore the concept of agency and intersectionality to interpret the interplay of forces that influence contraceptive use among Somali women refugees in Kakuma Camp indicated that there exist some beliefs that Islam is against condom use [20]. With the multireligious nature of the nationalities in the camp, it was useful to study more on the relationship between unwanted pregnancy and religion in Kakuma.

A study in Thailand attributed occurrence of adolescent pregnancy to a host of health system factors, including the availability of contraceptives and/or non-use of contraceptives alongside difficult access to health care and lack of sexual and reproductive health education which impacts SRH knowledge [21]. These findings highlight the vulnerability of refugee girls to unwanted pregnancies in refugee settings. It was useful to study how access to information, commodities and services affect adolescent pregnancy in Kakuma.

Unmet need for contraception remains an important contributor to unwanted pregnancy. In Kakuma, the contraceptive prevalence rate is approximately 45%, suggesting persistent gaps in access to and use of family planning services. Continued refugee arrivals from settings with weak health systems may further affect knowledge and practices related to contraception and safer sex. Despite ongoing sexual and reproductive health services provided by humanitarian agencies, unwanted pregnancy remains a concern. Although studies in other refugee settings have identified demographic, social, and health-system factors associated with unwanted pregnancy, these relationships have not been adequately examined in Kakuma Refugee Camp. This study therefore estimated the prevalence of unwanted pregnancy and examined socio-demographic factors associated with unwanted pregnancy among adolescent girls in Kakuma Refugee Camp.

## Materials and Methods

### Study site and population

This cross-sectional study was conducted at the Kakuma refugee camp which is located in North-Western Kenya in Turkana County in Kenya [22]. Kakuma refugee camp has an estimated 203,680 refugees of which approximately 12% represents adolescent girls aged 12-18 and it is the second largest refugee camp in Kenya with population diversity of over 10 nationalities [2]. The recruitment started on 1^st^ June 2026-31^st^ July 2026.

### Study population

The study population comprised approximately 16,859 UNHCR-registered adolescent girls aged 12–18 years residing in Kakuma Refugee Camp [2]. The sample size comprised 431 adolescent girls who were recruited through a multi-stage sampling approach to ensure a representative and systematic selection of study participants among refugee adolescent girls in Kakuma Refugee Camp, Turkana County.

### Inclusion and exclusion criteria

Those included were adolescent girls aged 12–18 years who were registered with UNHCR and who provided informed assent, with parental or guardian consent obtained where required. Exclusion criteria were adolescent girls who were severely ill at the time of data collection and unable to participate in the interview; those who were unable to provide informed assent or consent and for whom parental or guardian consent was not available where required; and those with severe cognitive impairment that precluded meaningful participation in the study.

### Sample Size determination

The minimum sample size was calculated using Yamane’s finite-population formula, n = N/[1 + N(e²)], where N was the estimated population of 16,859 registered adolescent girls and e was the desired precision of 0.05. This produced a minimum sample of 392 participants. After allowing 10% for non-response, the target sample was 431 adolescent girls.

### Sampling and participant recruitment

A multistage sampling procedure was used to recruit adolescent girls aged 12–18 years from Kakuma Refugee Camp. In the first stage, the four camp sections—Kakuma 1, Kakuma 2, Kakuma 3, and Kakuma 4 were treated as primary sampling units. Within each section, participants were stratified by zone and block to account for differences in population distribution.

The sample was allocated across the four camp sections using probability proportional to size, based on the population of eligible adolescent girls in each section. The UNHCR household registry was then used to identify households containing potentially eligible participants. Following household listing and eligibility screening, adolescent girls were selected using simple random sampling. The allocation is presented in Table 1.

**Table 1.** Allocation of the study sample across Kakuma camp sections.

|  | <u>Sub-Camp</u> | <u>12 to 18 years</u> | <u>Percentage</u> | <u>Proportionate Sampled girls of 12-18 years</u> | <u>Percentage</u> |
| --- | --- | --- | --- | --- | --- |
| 1 | Kakuma 1 | 6724 | 40% | 172 | 40% |
| 2 | Kakuma 2 | 2364 | 14% | 60 | 14% |
| 3 | Kakuma 3 | 4886 | 29% | 125 | 29% |
| 4 | Kakuma 4 | 2885 | 17% | 73 | 17% |
|  | Total | 16859 | 100% | 430 | 100% |

Source: Kakuma Census Report 2024

When a selected household had no eligible participant available at the time of data collection, the study team proceeded to the next household according to the predetermined sampling route. Written informed consent was obtained from participants aged 18 years. For participants younger than 18 years, written consent was obtained from a parent or legally authorized guardian, together with the participant’s assent.

For the qualitative component, four health workers (one from each camp section) were purposively selected for key informant interviews based on their experience providing services to adolescents. One focus group discussion was conducted with adolescent girls recruited from across the four camp sections. The interviews and focus group discussion were conducted in English or Kiswahili, digitally recorded with participants’ permission, transcribed verbatim, and translated into English where necessary. The transcripts were subsequently analysed thematically.

### Data collection instruments

Quantitative data were collected using a structured questionnaire administered to participating adolescent girls. The questionnaire covered three domains: sociodemographic characteristics, sexual and reproductive health factors, and contextual or moderating factors potentially associated with unwanted pregnancy.

Qualitative data were collected using semi-structured interview guides. The key informant guide explored health workers’ perspectives on demographic, social, and sexual and reproductive health-service factors associated with unwanted pregnancy in the refugee camp setting. A corresponding discussion guide was used for the focus group with adolescent girls.

### Pilot testing, validity, and reliability

The instruments were piloted in Kalobeyei Refugee Settlement among participants equivalent to approximately 10% of the intended study sample. Feedback from the pilot was used to clarify ambiguous questions, refine instructions and sequencing, and improve data-collection procedures. The revised instruments were reviewed by the university supervisors for content relevance and alignment with the study objectives. Questionnaire stability was assessed by administering the instrument to the same pilot participants twice, two weeks apart. Internal consistency was assessed using Cronbach’s alpha, with a coefficient of at least 0.70 prespecified as acceptable. Pilot data were excluded from the final analysis.

### Ethical considerations

Ethical approval for this study was obtained from the Maseno University Scientific and Ethics Review Committee (MUSERC; approval no. MUERC/809/19, reference no. MSU/DRPI/MUERC/00809/19, approved on 21 February 2025). Authorization to conduct the research was granted by the National Commission for Science, Technology and Innovation (NACOSTI; research licence no. NACOSTI/P/25/416850, Ref. No. 272744, issued on 20 March 2025). Permission to conduct the study in Kakuma Refugee Camp was also obtained from the relevant camp administration through the United Nations High Commissioner for Refugees (UNHCR) and the International Rescue Committee (IRC). The study was conducted in accordance with the ethical principles of the Declaration of Helsinki and applicable national and institutional ethical guidelines.

Written informed consent was obtained from all adult participants before participation. For participants younger than 18 years, written assent was obtained together with written informed consent from a parent or legally authorized guardian, as applicable and in accordance with the protocol approved by MUSERC. Participants received adequate information about the study, including its purpose, procedures, potential risks and benefits, confidentiality safeguards, and their right to decline or withdraw without penalty. Participation was voluntary, and participants’ privacy, confidentiality, and anonymity were maintained throughout the research.

### Outcome and covariate measurement

The variables on the socio-demographic factors were developed specifically for this study. The study questionnaire is provided in S1 Appendix. The questions on pregnancy intention were adopted from the Kenya Demographic and Health Survey (KDHS) method of Pregnancy intention measurement. In the KDHS woman’s questionnaire, pregnancy intention is measured by respondent’s answer to the following question: “When you got pregnant with (NAME), did you want to get pregnant at that time, did you want to wait until later, or did you not want to have any child at all?”. Responses indicating that the pregnancy was wanted at the time or wanted later were classified as no unwanted pregnancy while responses indicating that no child was wanted at all were classified as unwanted pregnancy.

### Data analysis

Data were entered into Excel 2010, pre-coded then analysed using IBM SPSS Version 27.0. Quantitative data collected using the questionnaires were checked for completeness, coded, entered and analyzed using SPSS version 27. Both descriptive and inferential statistics were used. For descriptive statistics, tables and percentages were used to summarize the findings. The chi-square test of independence was used to assess differences in the proportions of adolescents who reported unwanted pregnancy and those who did not, and to determine associations between sociodemographic factors and unwanted pregnancy among adolescents. Statistically significant associations were further subjected to logistic regression. Statistical significance was set at p≤0.05. Qualitative data from the one FGD and 4 key informant interviews were subjected to manual thematic content analysis. Audio recordings were transcribed verbatim and, where interviews were conducted in Kiswahili, translated into English. The transcripts were reviewed for accuracy, read repeatedly for familiarization, and manually coded. Related codes were grouped into categories, themes, and subthemes. Findings from the FGD and KIIs were compared to identify areas of convergence and divergence and were subsequently triangulated with the quantitative findings.

## Results

### Demographic characteristics of adolescent girls at Kakuma refugee camp

A total of 431 adolescent girls participated in the study. Two adolescent girls declined to provide assent and were replaced in accordance with the sampling procedure; ultimately, all 431 eligible adolescent girls were included in the final sample. The age of respondents ranged from 12 to 18 years: 22.5% were aged 12–14 years and 77.5% were aged 15–18 years. Regarding marital status, 19.2% were married and 80.8% were single/divorced/separated. Participants were distributed across Kakuma 1 (39.9%), Kakuma 2 (13.9%), Kakuma 3 (29.0%), and Kakuma 4 (17.2%). Overall, 67.5% were in school and 32.5% were out of school. Most participants were Christian (74.7%), followed by Muslim (23.9%), African Traditional Religion (0.9%), and other faiths (0.2%). Household heads included mothers, fathers, spouses, and other relatives. With respect to pregnancy intention, 28.7% of participants reported having experienced an unwanted pregnancy, while 71.3% did not. Place of birth varied considerably, and the main reasons for moving to Kenya included joining family, marriage, work, schooling, escaping insecurity/war/drought, and other reasons. Participant characteristics are summarized in Table 2.

**Table 2.**
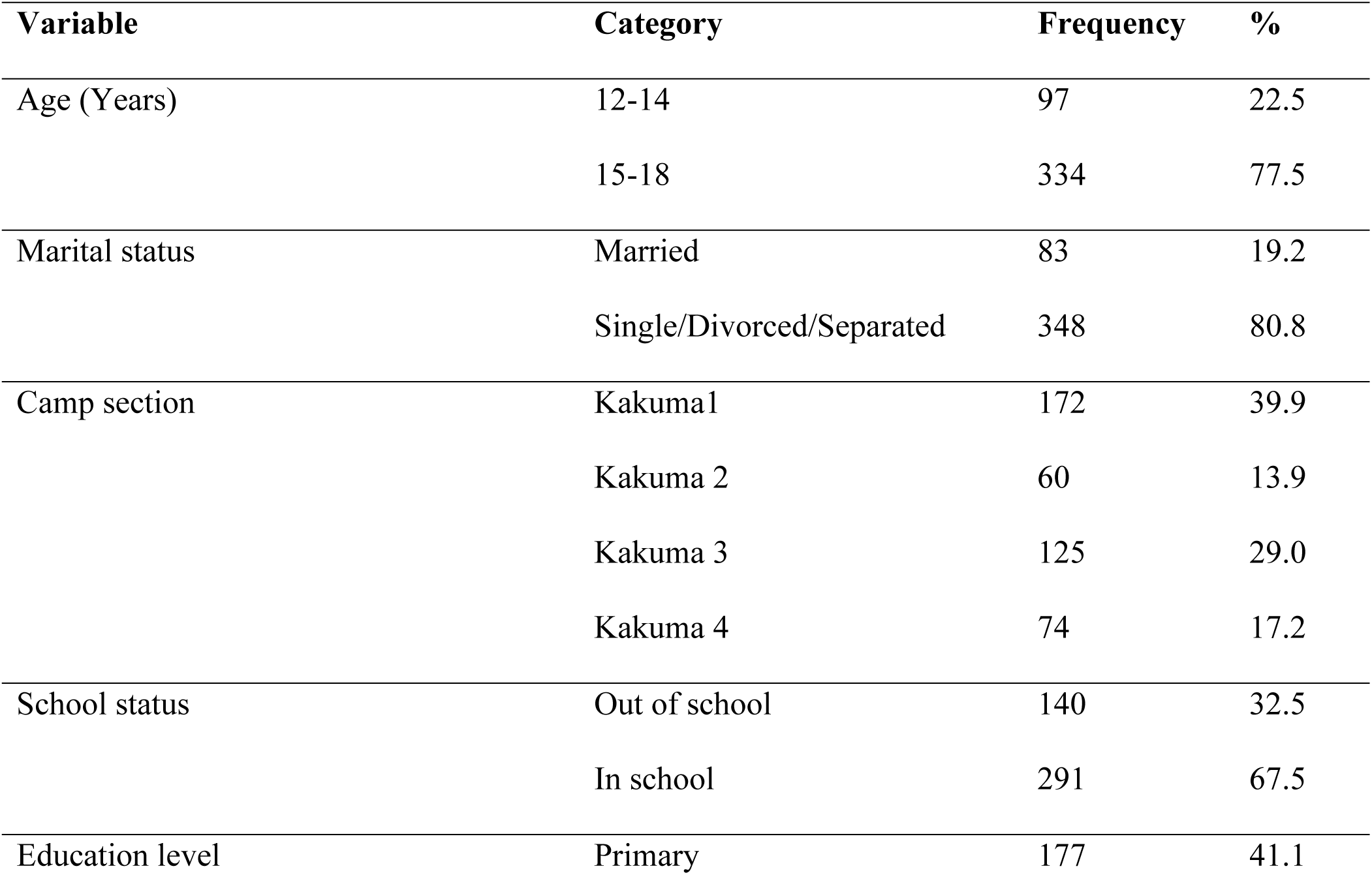
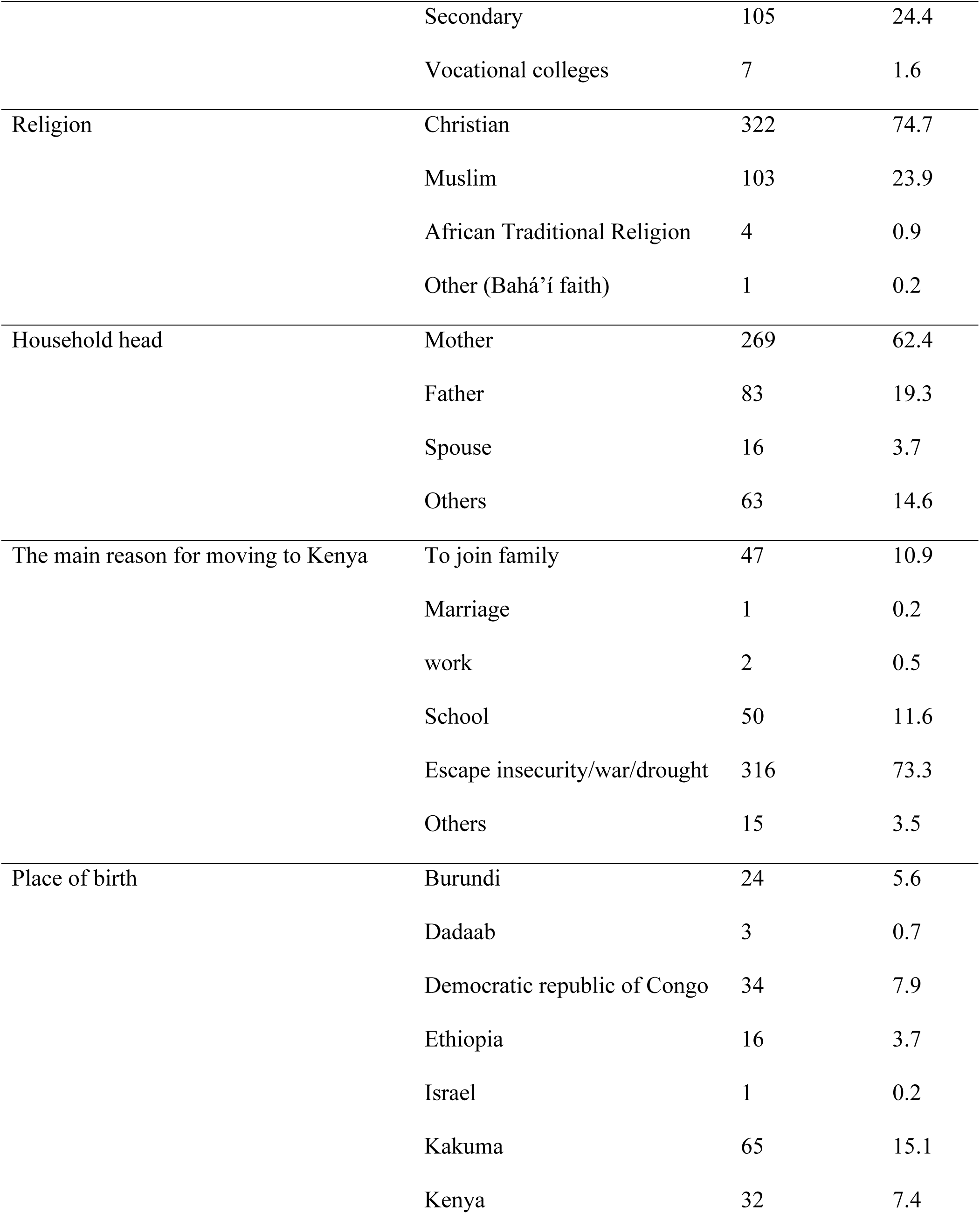
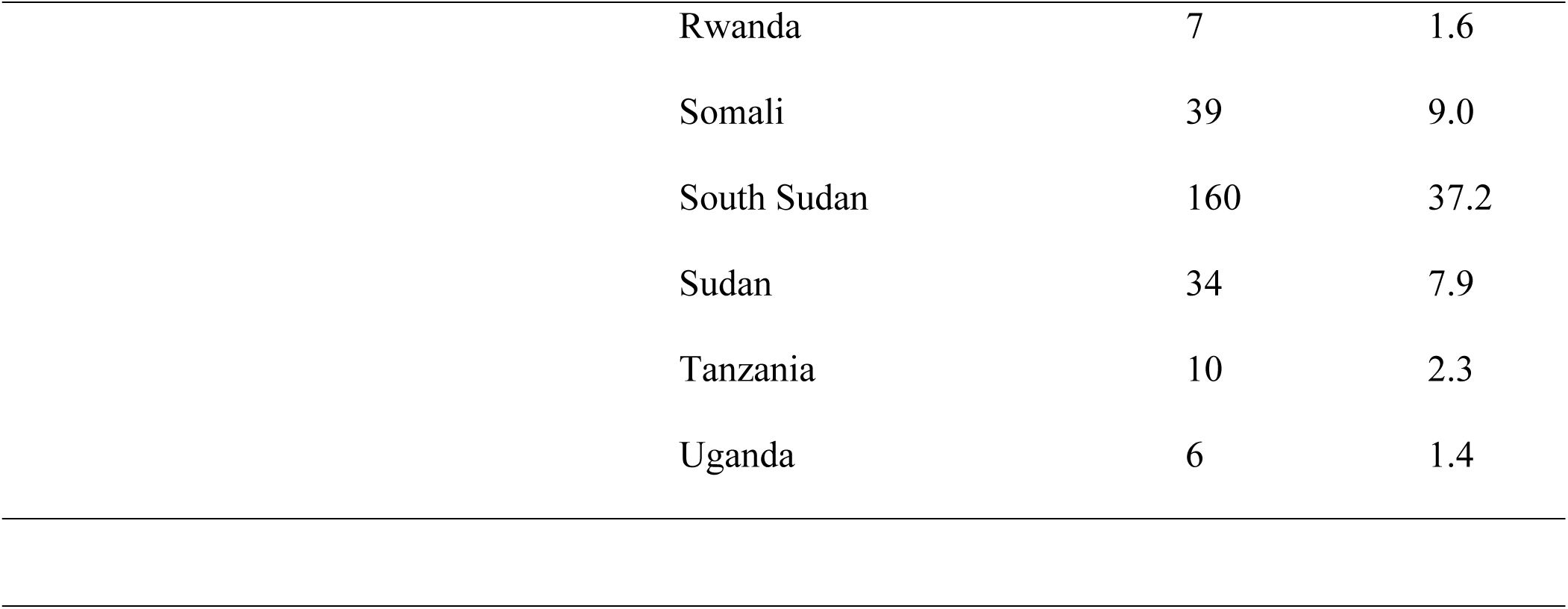
Characteristics of the study participants.

| Variable | Category | Frequency | % |
| --- | --- | --- | --- |
| Age (Years) | 12-14 | 97 | 22.5 |
|  | 15-18 | 334 | 77.5 |
| Marital status | Married | 83 | 19.2 |
|  | Single/Divorced/Separated | 348 | 80.8 |
| Camp section | Kakuma1 | 172 | 39.9 |
|  | Kakuma 2 | 60 | 13.9 |
|  | Kakuma 3 | 125 | 29.0 |
|  | Kakuma 4 | 74 | 17.2 |
| School status | Out of school | 140 | 32.5 |
|  | In school | 291 | 67.5 |
| Education level | Primary | 177 | 41.1 |

**Table 2. Characteristics of the study participants**
|  |  |  |  |
| --- | --- | --- | --- |
|  | Secondary | 105 | 24.4 |
|  | Vocational colleges | 7 | 1.6 |
| Religion | Christian | 322 | 74.7 |
|  | Muslim | 103 | 23.9 |
|  | African Traditional Religion | 4 | 0.9 |
|  | Other (Bahá'í faith) | 1 | 0.2 |
| Household head | Mother | 269 | 62.4 |
|  | Father | 83 | 19.3 |
|  | Spouse | 16 | 3.7 |
|  | Others | 63 | 14.6 |
| The main reason for moving to Kenya | To join family | 47 | 10.9 |
|  | Marriage | 1 | 0.2 |
|  | work | 2 | 0.5 |
|  | School | 50 | 11.6 |
|  | Escape insecurity/war/drought | 316 | 73.3 |
|  | Others | 15 | 3.5 |
| Place of birth | Burundi | 24 | 5.6 |
|  | Dadaab | 3 | 0.7 |
|  | Democratic republic of Congo | 34 | 7.9 |
|  | Ethiopia | 16 | 3.7 |
|  | Israel | 1 | 0.2 |
|  | Kakuma | 65 | 15.1 |
|  | Kenya | 32 | 7.4 |

**Table 2. Characteristics of the study participants**
|  |  |  |
| --- | --- | --- |
| Rwanda | 7 | 1.6 |
| Somali | 39 | 9.0 |
| South Sudan | 160 | 37.2 |
| Sudan | 34 | 7.9 |
| Tanzania | 10 | 2.3 |
| Uganda | 6 | 1.4 |

### Prevalence of unwanted pregnancies among adolescent girls in Kakuma refugee camp Quantitative findings

Pregnancy intention was assessed using the Kenya Demographic and Health Survey (KDHS) pregnancy-intention question. Based on participants’ reported pregnancy intention, 124 adolescents were classified as having experienced an unwanted pregnancy. The prevalence of unwanted pregnancy in the study population was therefore 28.7% (124/431). The prevalence of unwanted pregnancy varied across the four camp sections. In Kakuma 1, 20.9% (36/172) of participants were classified as having experienced an unwanted pregnancy; in Kakuma 2, 56.7% (34/60); in Kakuma 3, 11.2% (14/125); and in Kakuma 4, 54.1% (40/74).

### Prevalence of unwanted pregnancy by country of origin

The prevalence of unwanted pregnancy also varied by country of origin. Among adolescents from South Sudan, 41.9% (67/160) reported having experienced an unwanted pregnancy, compared with 21.0% (57/271) among adolescents from other countries of origin. These findings are presented in Table 3.

**Table 3.**
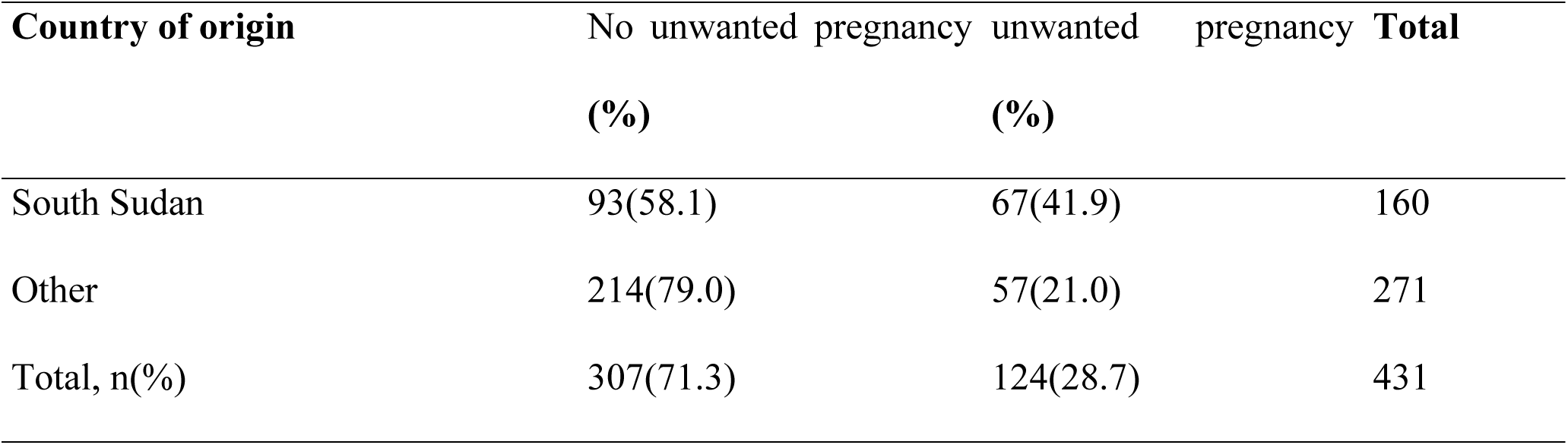
Prevalence of unwanted pregnancy by country of origin.

### Qualitative findings

Qualitative data from one focus group discussion (FGD) and four key informant interviews (KIIs) were analyzed thematically. The findings were organized around participants’ understanding of unwanted pregnancy and the factors contributing to unwanted pregnancy among adolescent girls in Kakuma Refugee Camp.

### Theme 1: Unwanted pregnancy as an unplanned and socially disruptive experience

Participants generally understood unwanted pregnancy as a pregnancy that occurs unexpectedly and is neither intended nor socially accepted. Beyond being unplanned, participants described such pregnancies as disruptive to an adolescent girl’s life and potentially associated with stigma and negative social consequences. This understanding suggests that unwanted pregnancy was perceived not only in terms of reproductive intention but also in relation to its effects on an adolescent girl’s social position and future prospects.

There was convergence between the FGD and KIIs in recognizing unwanted adolescent pregnancy as an important concern within the refugee camp. However, while FGD participants emphasized the personal and social consequences of an unwanted pregnancy, key informants placed greater emphasis on the wider health consequences and inadequate access to sexual and reproductive health information.

> *“It’s a pregnancy that’s abrupt/sudden, not accepted by society, viewed as a curse, one that disrupted your normal life as a child or a girl.”* FGD participant
>
> *“It’s more prevalent if not given safety SRH education and it leads to increased criminal abortions that led to high mortality rates among teenagers.”* Key informant 1

### Theme 2: Economic vulnerability and unmet basic needs as drivers of unwanted pregnancy

Economic hardship emerged as a major factor contributing to unwanted pregnancy among adolescent girls. Participants described inadequate household resources, inability to meet education related costs, and pressure to provide for siblings as circumstances that could increase adolescents’ vulnerability. Material and social aspirations, including the desire to obtain clothing and other items associated with fitting in among peers, were also reported as factors influencing relationships with potential sexual partners.

The FGD and Key informant interviews converged in identifying economic vulnerability as an important underlying driver of adolescent pregnancy. FGD participants provided more detailed accounts of specific household and personal needs, including food, school fees, school supplies and responsibility for siblings. Key informants similarly described the broader socioeconomic vulnerability experienced by adolescent girls in the camp and indicated that some girls entered relationships as a means of obtaining financial or material support.

> *“It’s because of inadequate resources, i.e., feeding, fees and school items… As the head of household, you need to provide for the siblings.”* FGD participant
>
> *“Vulnerability in the camp leads to adolescent girls getting unwanted pregnancy and led the adolescents to engage in relationships so that they can get some income.”* Key informant 2

### Socio-demographic factors associated with unwanted pregnancies among adolescent refugee girls in Kakuma refugee camp

#### Quantitative findings

Age was significantly associated with unwanted pregnancy (χ² = 28.376, p < 0.001), with 35.0% of adolescents aged 15–18 years reporting an unwanted pregnancy compared with 7.2% of those aged 12–14 years. Marital status was also significantly associated with unwanted pregnancy (χ² = 45.708, p < 0.001): 59.0% of married adolescents reported an unwanted pregnancy compared with 21.6% of single/divorced/separated adolescents. School attendance was significantly associated with unwanted pregnancy (χ² = 20.079, p < 0.001), as was education level (χ² = 4.929, p = 0.026). Religion (χ² = 12.054, p < 0.001), household headship (χ² = 12.619, p = 0.013), and camp section (χ² = 69.858, p < 0.001) were also significantly associated with unwanted pregnancy. Current enrolment in a cash transfer/food assistance programme was associated with unwanted pregnancy (χ² = 7.875, p = 0.005), whereas income-generating activity, ever having been enrolled in a cash transfer/food assistance programme, household food deprivation, parental status, and reason for coming to the camp were not statistically significant. Bivariable associations are presented in Table 4.

**Table 4.**
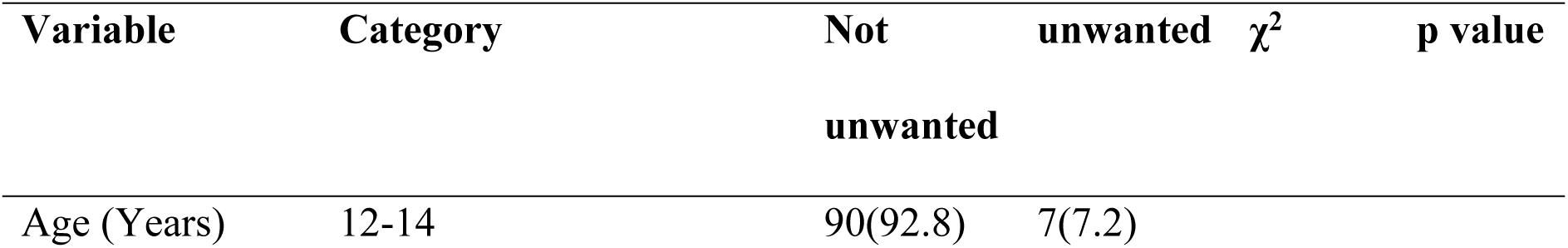
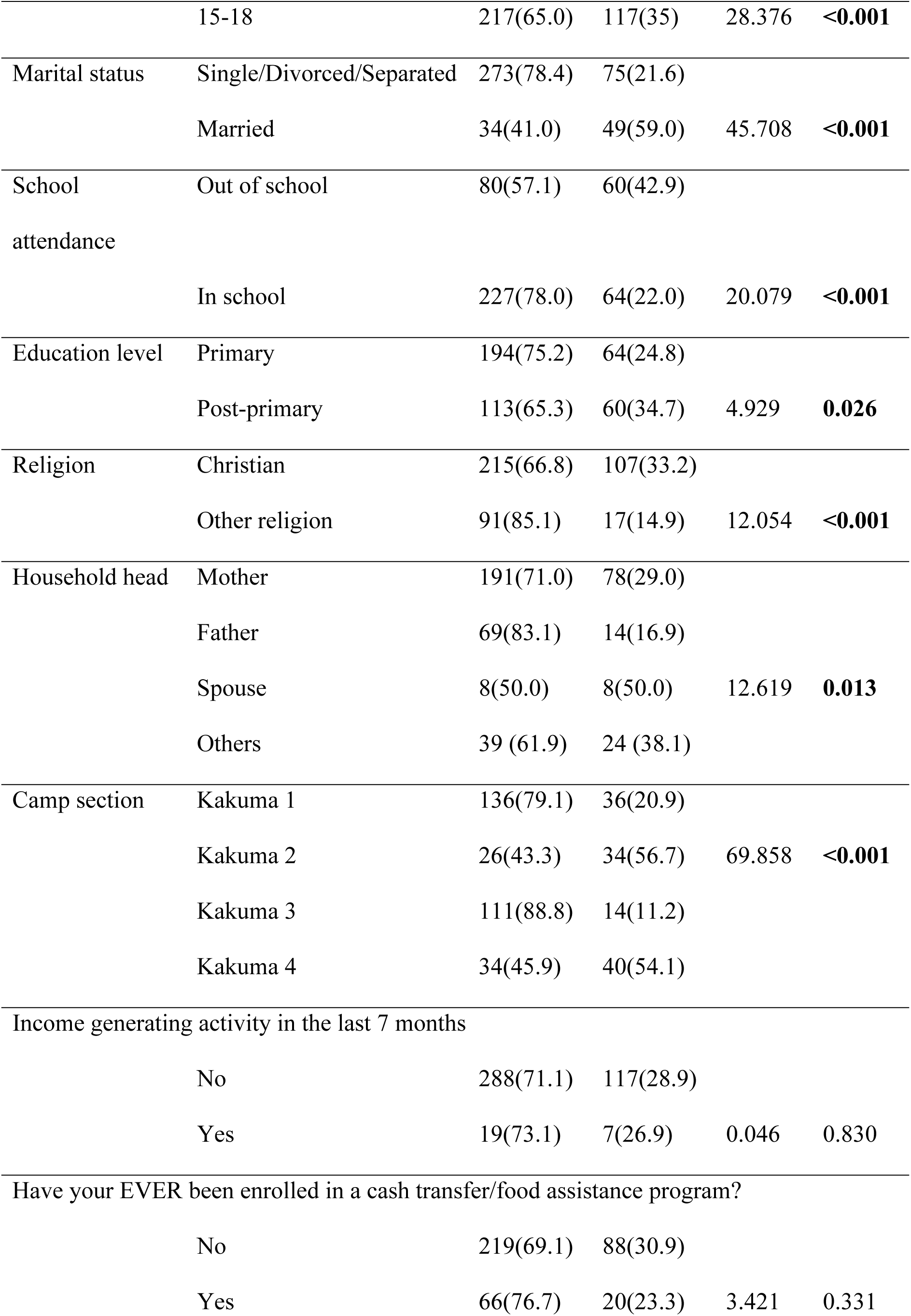
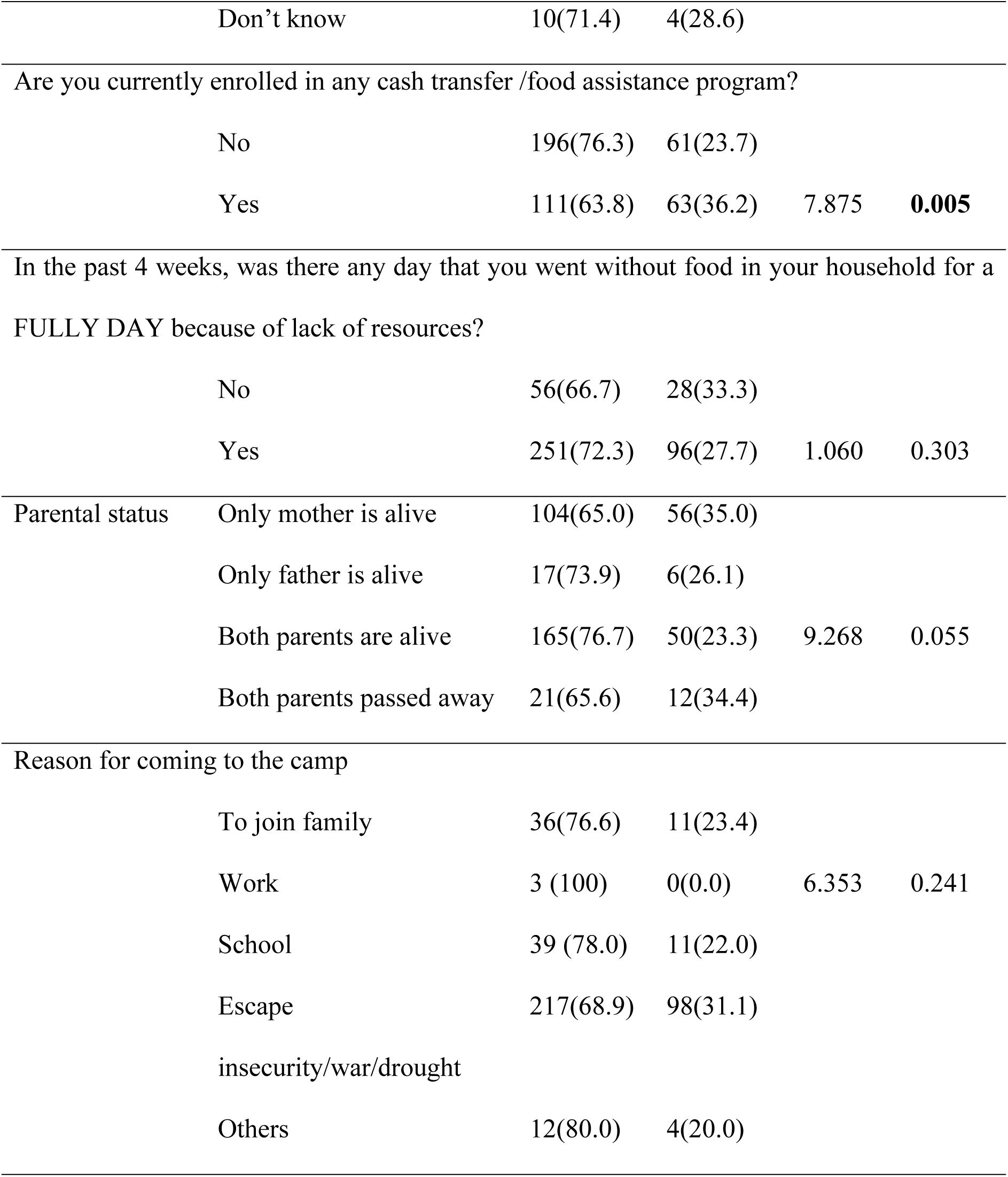
Bivariable associations with unwanted pregnancy.

### Association between socio-demographic factors and unwanted pregnancy among adolescents

Bivariable and multivariable logistic regression further characterized the associations presented in Tables 4 and 5. Adolescents aged 12–14 years had substantially lower crude odds of unwanted pregnancy than those aged 15–18 years (COR = 0.144; 95% CI: 0.065–0.321; p < 0.001); after adjustment, the association was attenuated and borderline statistically significant (AOR = 0.348; 95% CI: 0.121–1.000; p = 0.050). Married adolescents had markedly higher odds of unwanted pregnancy than single/divorced/separated adolescents in both crude (COR = 5.22; 95% CI: 3.149–8.675; p < 0.001) and adjusted analyses (AOR = 5.108; 95% CI: 3.065–8.512; p = 0.001). Adolescents who were out of school had higher crude odds than those in school (COR = 2.660; 95% CI: 1.722–4.109; p < 0.001); no adjusted estimate was presented for school attendance. Primary education was associated with lower crude odds than post-primary education (COR = 0.621; 95% CI: 0.408–0.947; p = 0.027), but the adjusted association was not statistically significant (AOR = 0.660; 95% CI: 0.419–1.038; p = 0.072). Christian adolescents had higher adjusted odds than adolescents of other religions (AOR = 3.010; 95% CI: 1.087–8.337; p = 0.034). For household headship, the lower crude odds observed among adolescents in father-headed households (COR = 0.330; 95% CI: 0.153– 0.710; p = 0.005) were attenuated after adjustment (AOR = 0.869; 95% CI: 0.271–3.328; p = 0.838). Using Kakuma 4 as the reference, adolescents in Kakuma 1 (AOR = 0.168; 95% CI: 0.059–0.483; p < 0.001) and Kakuma 3 (AOR = 0.073; 95% CI: 0.021–0.248; p < 0.001) had lower adjusted odds of unwanted pregnancy, while the association for Kakuma 2 was not statistically significant (AOR = 0.560; 95% CI: 0.158–1.985; p = 0.369).

**Table 5.**
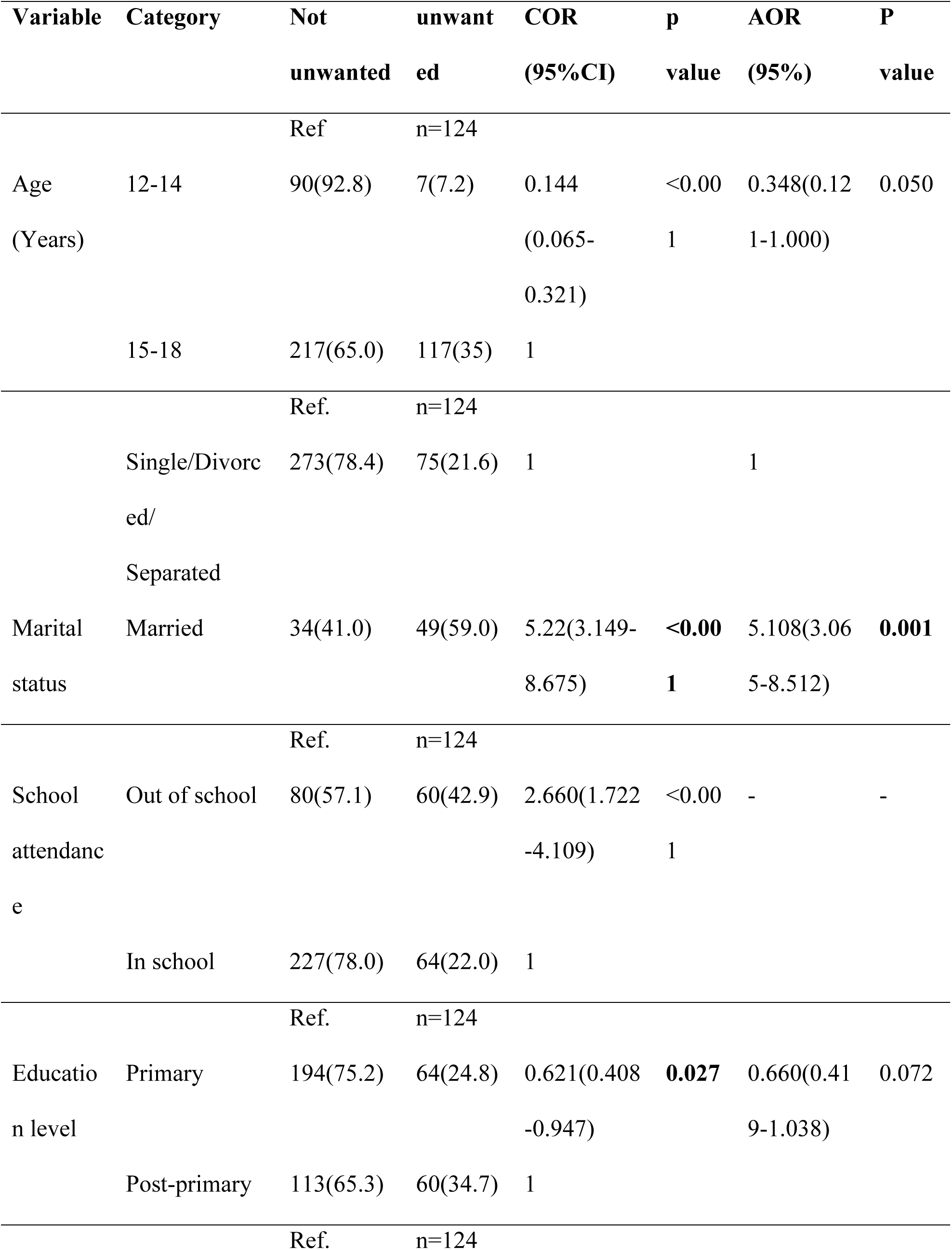
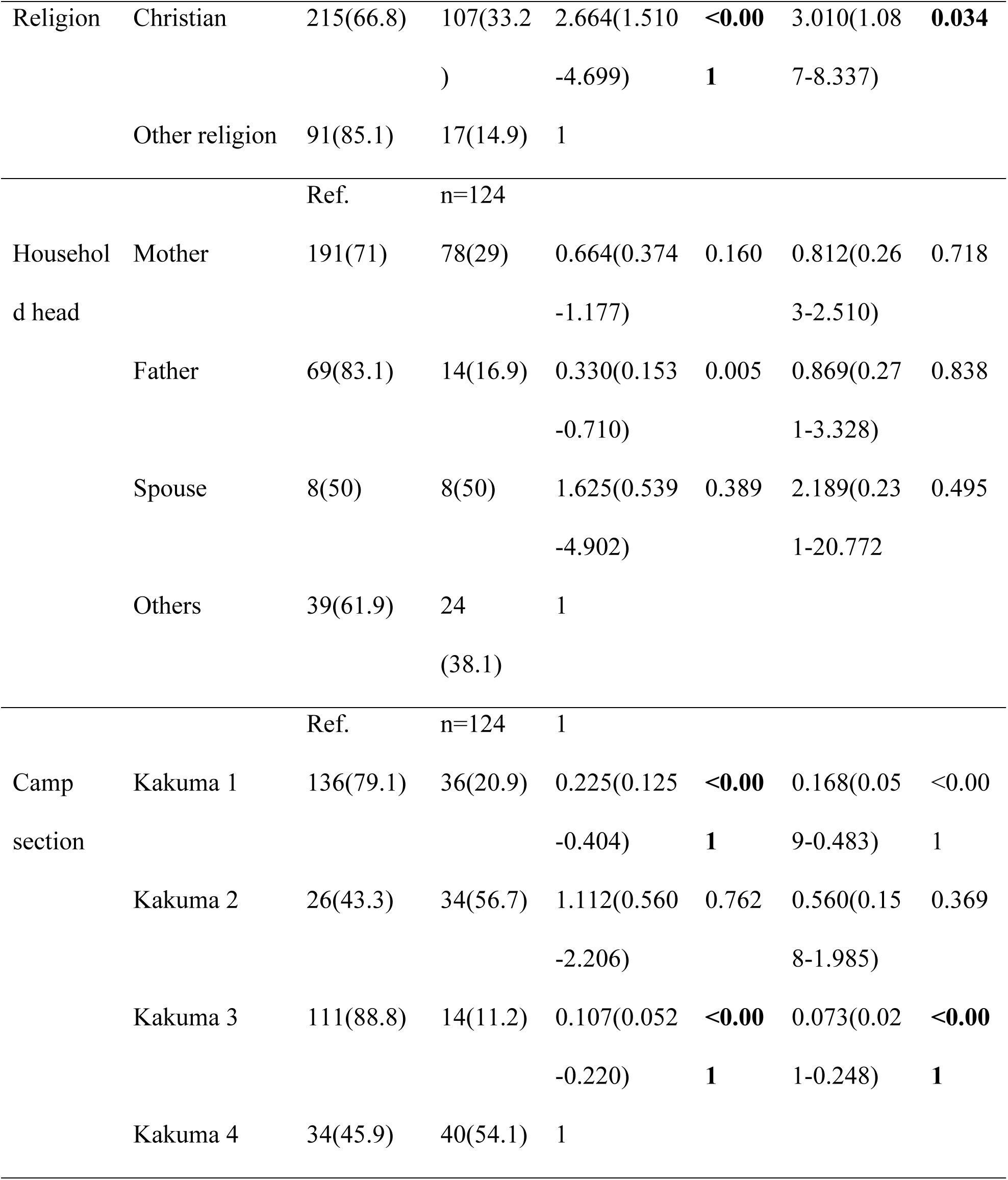
Logistic regression results.

### Qualitative findings

#### Theme 3: Perceptions of the appropriate age for sexual initiation

Participants expressed the view that sexual activity should ideally begin in early adulthood, particularly after completion of schooling and when an individual is considered physically and socially prepared for marriage and childbearing. These perspectives suggest that perceptions of appropriate sexual initiation were closely linked to educational attainment, physical maturity, marriage, and readiness for parenthood rather than age alone.

The available data in this subtheme are drawn mainly from the FGD. Participants converged around the view that sexual initiation should be delayed until a young woman has completed school and reached an age considered socially appropriate for marriage and childbearing. No corresponding informant interview perspective is presented in the available excerpt, limiting assessment of convergence or divergence between adolescents and key informants.

> *“20 years and above because at that time one is done with school and the body is ready to have the baby.”* — FGD participant
>
> *“It’s a societally appropriate age when one should get married and have a child.”* — FGD participant

#### Theme 4: Religion as a platform for abstinence-focused pregnancy prevention

Religion emerged as an important social institution through which adolescents received messages related to prevention of unwanted pregnancy. Participants indicated that direct discussion of unwanted pregnancy within religious settings was limited; however, churches provided opportunities to engage young people on sexual behaviour, particularly during holiday youth activities. The dominant prevention message reported was abstinence.

The available evidence for this theme comes from the FGD and therefore does not permit direct comparison with KII perspectives. The finding nevertheless suggests a distinction between the limited direct discussion of unwanted pregnancy and the more prominent use of abstinence-focused messaging within faith-based youth activities.

> *“Unwanted pregnancy is a topic that’s less spoken about, but during holiday youth activities in church they mostly focus on abstinence.”* — FGD participant

## Discussion

### Prevalence and contextual differences

This study found a high prevalence of unwanted pregnancy among adolescent girls in Kakuma Refugee Camp, with 28.7% of participants reporting having experienced an unwanted pregnancy based on the KDHS pregnancy-intention measure [23]. This relatively high prevalence may reflect the multiple sexual, reproductive, social and economic vulnerabilities experienced by adolescent girls in humanitarian settings. Prevalence of adolescent pregnancy in a refugee camp has been shown to vary considerably from 21% to 41.2% depending on regions [24]. This finding is consistent with findings from Benin [25], eastern Ethiopia [26] and in Kule refugee camp, Gambella Ethiopia [5]. The broader SRH vulnerability reflected in this finding is also consistent with evidence of suboptimal contraceptive use among female refugee adolescents in northern Uganda [27].

The similarity in prevalence observed between the current study and findings from other refugee settings may reflect comparable socio-demographic and socio-economic conditions, as well as the heightened vulnerability of adolescent girls to sexual and reproductive health challenges in humanitarian contexts. Such vulnerabilities may increase exposure to unprotected sexual activity and consequently the risk of pregnancy [27]. For example, a study conducted in northern Uganda reported that more than one-quarter of adolescent girls in a refugee camp were sexually active [28]. However, the prevalence observed in the current study was lower than that reported in studies from Malawi [15], Chad [29], Cameroon [30], and parts of Ethiopia [31]. These differences may be explained by variations in study design, sample size, age distribution of participants, study period, and recruitment setting. For instance, Ahinkorah et al. [29] conducted a multicountry study across sub-Saharan Africa between 2010 and 2018, while studies by Kaphagawani et al. [15] and Donatus et al. [30] were clinic-based and therefore more likely to include adolescents who were already pregnant and attending antenatal services. Such methodological differences limit direct comparison of prevalence estimates across studies.

### Age-related factors

Age was strongly associated with unwanted pregnancy at the crude level. Adolescents aged 12–14 years had substantially lower crude odds than those aged 15–18 years, while the adjusted estimate was attenuated and reached the conventional threshold only marginally (AOR = 0.348; 95% CI: 0.121–1.000; p = 0.050). The direction of effect remains consistent with studies among refugee populations in Uganda and Ethiopia by Okello et al. [24], Ivanova et al. [35], and Adhena and Fikre [5], as well as a community-based study in Uganda by Akol et al. [36]. Older adolescents may have greater exposure to sexual relationships, earlier unions, and other social circumstances that increase the likelihood of pregnancy, as well as a longer cumulative period of sexual exposure. The attenuated adjusted association in this study suggests that part of the age-related difference may overlap with marital, religious, educational, or contextual factors. These findings nevertheless support ensuring that age-appropriate sexual and reproductive health information and contraceptive services are accessible before and throughout later adolescence.

### Marital and school status

Marital status was one of the clearest independent correlates of unwanted pregnancy. Married adolescents had approximately five-fold higher adjusted odds of unwanted pregnancy compared with single/divorced/separated adolescents (AOR = 5.108; 95% CI: 3.065–8.512), consistent with findings from Kule Refugee Settlement in Ethiopia [5]. This association may reflect greater frequency of sexual activity within marriage, social expectations regarding childbearing, and limited autonomy in reproductive decision-making. In some settings, married adolescent girls may face pressure to demonstrate fertility soon after marriage, while access to family planning may be constrained by partner or family opposition and prevailing cultural beliefs [4]. At the same time, the relationship between marriage and pregnancy may be bidirectional because pregnancy itself may precede or precipitate marriage in some circumstances [38]. Given the cross-sectional design, the temporal sequence between marital status and unwanted pregnancy cannot be established.

School attendance was significantly associated with unwanted pregnancy in the bivariate analysis: adolescents who were out of school had higher crude odds than those attending school (COR = 2.660; 95% CI: 1.722–4.109). However, Table 5 does not present an adjusted estimate for school attendance, and the finding should therefore not be interpreted as an independent adjusted determinant. The crude association is nevertheless consistent with previous studies showing that school participation may be protective against adolescent pregnancy [24, 39, 40]. The qualitative findings indicated inability to meet school-related costs and economic vulnerability may place girls in relationships for material support. School attendance may provide structured daily engagement, access to sexual and reproductive health information, peer and adult support, and stronger expectations regarding future education and employment. Conversely, adolescents who are out of school may experience greater exposure to early marriage, economic dependency, transactional relationships, and other vulnerabilities. Pregnancy itself may also lead to school interruption or dropout, so the observed relationship should not be interpreted as demonstrating temporal causation.

### Contextual and camp-related factors

Camp section remained an important independent correlate after adjustment. Compared with adolescents residing in Kakuma 4, those in Kakuma 1 (AOR = 0.168; 95% CI: 0.059–0.483) and Kakuma 3 (AOR = 0.073; 95% CI: 0.021–0.248) had substantially lower adjusted odds of unwanted pregnancy, whereas the adjusted odds in Kakuma 2 did not differ significantly from Kakuma 4. These findings are consistent with the marked descriptive variation across sections, with unwanted pregnancy prevalence highest in Kakuma 2 (56.7%) and Kakuma 4 (54.1%) and lowest in Kakuma 3 (11.2%). Differences between camp sections may reflect variation in population composition, country of origin, socioeconomic conditions, educational opportunities, safety, social networks, and access to sexual and reproductive health information and services. In the current study, camp sections with higher unwanted pregnancy prevalence included a larger proportion of adolescents originating from South Sudan, and adolescents from South Sudan also accounted for a substantial proportion of unwanted pregnancies. Camp location may therefore partly capture underlying cultural, socioeconomic, and displacement-related characteristics rather than acting as a causal factor in itself.

The variation observed across Kakuma camp sections should therefore be interpreted as a possible marker of unequal exposure to social and structural conditions. Differences in the availability or accessibility of adolescent-responsive services, contraceptive information, schooling, protection mechanisms, and safe spaces may contribute to variation in pregnancy risk across camp sections. These findings suggest the need for more geographically targeted assessment within the camp to determine whether particular sections experience greater service gaps or protection vulnerabilities.

Religion also remained independently associated with unwanted pregnancy: Christian adolescents had higher adjusted odds than adolescents of other religions (AOR = 3.010; 95% CI: 1.087–8.337; p = 0.034). This association should be interpreted cautiously because religion may operate indirectly through norms surrounding sexuality, marriage, fertility, contraceptive use, and care-seeking rather than through religious affiliation itself [41]. Household headship, by contrast, did not remain independently associated after adjustment. Although adolescents in father-headed households had lower crude odds of unwanted pregnancy, this association was attenuated in the multivariable model, suggesting confounding by other social or demographic characteristics. Education showed a similar pattern: the crude association between primary versus post-primary education was statistically significant, but the adjusted estimate was not. The FGD evidence suggestss that churches are important platforms for abstinence-focused messaging.

Overall, the findings suggest that unwanted pregnancy among adolescent girls in Kakuma Refugee Camp is shaped by an interaction between individual, social, and contextual factors. Married adolescents were a particularly high-risk group in the adjusted model, while religion and camp section also remained independently associated with unwanted pregnancy. Age showed strong crude differences but only borderline adjusted evidence, and school attendance was associated at the crude level without an adjusted estimate in the final table. Prevention strategies should therefore combine adolescent-responsive contraceptive information and services with engagement of married adolescents and their partners, school retention and re-entry support, and geographically targeted assessment of higher-burden camp sections.

### Limitations

This study has several limitations. Its cross-sectional design precludes determination of temporality or causality. Sensitive sexual and reproductive health information was self-reported and may therefore be affected by recall and social-desirability bias, although interviews were conducted privately by trained data collectors. Replacement of selected participants who declined or were unavailable may have introduced selection bias. The aggregation of categories with small cell counts, particularly for religion and marital status, may have obscured heterogeneity and produced imprecise estimates. Residual confounding is also possible. The qualitative component comprised one focus group discussion and four key informant interviews and may not have achieved thematic saturation. Finally, the findings are primarily generalizable to adolescent girls in Kakuma Refugee Camp and should be transferred to other refugee or humanitarian settings cautiously.

### Recommendations

The Government of Kenya, UNHCR, and implementing partners should strengthen adolescent-responsive sexual and reproductive health services in Kakuma, with particular attention to married adolescents, school retention and re-entry, and camp sections with higher observed burdens. Routine pregnancy data should be disaggregated by age, marital status, school attendance, and camp section to support locally targeted planning. The observed association with religion requires further investigation and should not be interpreted as causal. Longitudinal and multilevel studies are needed to clarify the social, service-access, and protection-related mechanisms underlying variation within the camp.

### Conclusions

Unwanted pregnancy remains an important public health concern among adolescent girls in Kakuma Refugee Camp, affecting 28.7% of participants. Married adolescents had substantially higher adjusted odds of unwanted pregnancy, Christian adolescents also had higher adjusted odds than adolescents of other religions, and marked independent differences were observed across camp sections, particularly for Kakuma 1 and Kakuma 3 relative to Kakuma 4. Age was strongly associated at the crude level but was only borderline statistically significant after adjustment, while school attendance and education level showed crude associations without statistically significant adjusted evidence in the results presented. These findings support differentiated SRH interventions that engage married adolescents, support school retention and re-entry, and target higher-burden or underserved camp sections. Further longitudinal research is needed to clarify causal pathways and mechanisms underlying within-camp variation.

## Data Availability

Data Availability Statement The datasets generated and analyzed during the current study are not publicly available because they contain sensitive information collected from adolescent refugee girls on pregnancy intention and reproductive health. Given the vulnerability of the study population and the Kakuma refugee setting, open access to individual level data could compromise participant privacy and confidentiality, including through deductive identification. De-identified data supporting the findings of this study may be made available from the corresponding author upon reasonable request, subject to approval by the relevant ethics review committee, institutional data sharing requirements, and any applicable permissions from the refugee camp/implementing authorities. Data access will be limited to the minimum anonymized dataset required to verify the results reported in this article and will be governed by appropriate data-use and confidentiality agreements.

## Acknowledgments

We thank the adolescent girls who participated in this study and the health workers who contributed to the qualitative interviews. We also acknowledge the Kakuma Refugee Camp administration, UNHCR, and the International Rescue Committee for facilitating the conduct of the study.

## Supporting information

S1 Appendix. Study questionnaire. Questionnaire used to collect quantitative data from adolescent girls aged 12–18 years in Kakuma Refugee Camp.

